# Low-pass Whole Genome Imputation Enables the Characterization of Polygenic Breast Cancer Risk in the Indigenous Arab Population

**DOI:** 10.1101/2022.12.07.22282785

**Authors:** Mohammed Al-Jumaan, Hoyin Chu, Abdullah Al-Sulaiman, Sabrina Y. Camp, Seunghun Han, Riaz Gillani, Yousef Al Marzooq, Fatmah Almulhim, Chittibabu Vatte, Areej Al Nemer, Afnan Almuhanna, Eliezer M Van Allen, Amein Al-Ali, Saud H AlDubayan

## Abstract

The indigenous Arab population has traditionally been underrepresented in cancer genomics studies, and as a result the polygenic risk landscape of breast cancer in the population remains elusive. Here we show by utilizing low-pass whole genome sequencing (lpWGS), we can accurately impute population-specific variants with high exome concordance (median dosage correlation: 0.9459, Interquartile range: 0.9410-0.9490) and construct breast cancer burden-sensitive polygenic risk scores (PRS) using publicly available resources. After adjusting the PRS to the Arab population, we found significant associations between PRS performance in risk prediction and first-degree relative breast cancer history prediction (Spearman rho=0.43, p = 0.03), where breast cancer patients in the top PRS decile are 5.53 (95% CI: 1.76-17.97, p = 0.003) times more likely to also have a first degree relative diagnosed with breast cancer compared to those in the middle deciles. In addition, we found evidence for the genetic liability threshold model of breast cancer where among patients with a family history of breast cancer, pathogenic rare variant carriers had significantly lower PRS than non-carriers (p = 0.0205, M.W.U.) while for non-carriers every standard deviation increase in PRS corresponded to 4.52 years (95% CI: 8.88-0.17, p = 0.042) earlier age of presentation. Overall, our study provides a viable strategy utilizing lpWGS to assess polygenic risk in an understudied population and took steps in addressing existing global health disparities.

## Introduction

Individuals from the Greater Middle Eastern (GME) regions are underrepresented in genomic studies, with less than 0.01% of the samples in Genome-wide Association Studies (GWAS) Catalog [1] and less than 0.8% of the samples in the Genome Aggregation Database (gnomAD)[2] reporting GME origin[3]. Moreover, GME populations have roughly a doubling rate of recessive Mendelian disease compared to their European counterpart[4] and are experiencing a growing burden of breast cancer [5]. By understanding the germline genetic risk landscape of breast cancer in GME populations, not only could we broaden our understanding of predisposing genetics in diverse populations, but we could also address growing health disparities by gaining clinically relevant insights. Recent progress with the Qatar Genome Programme, which sequenced over 6,000 Qatari subjects with diverse GME ancestry backgrounds has revealed significant differences in breast cancer polygenic risk score (PRS) distributions between cancer-free populations with different GME ancestry backgrounds [6].

These findings have highlighted the urgency of understanding the role of polygenic risk in the GME populations, and the further characterization of polygenic risk among breast cancer patients of GME ancestry and their relations to the clinical presentation of breast cancer is needed.

Low-pass whole genome sequencing (lpWGS), or WGS with an average sequencing depth of around 1.0x, has recently been proposed as a cost-effective alternative data modality to study genetic architectures in understudied populations. Compared to the traditional genotyping arrays, lpWGS has reduced genetic variant ascertainment bias and has been shown to be sensitive to population-specific novel variants [7]. In addition, lpWGS has also been shown to outperform genotype arrays in imputation performance and statistical power [8–10]. Given these advantages, lpWGS appears as an attractive option to understand the polygenic architecture of breast cancer in GME populations, but its accuracy and utility beyond risk prediction have yet to be systematically evaluated in a clinical setting.

In this multi-center study, we collected blood samples from 220 female breast cancer patients from the indigenous Arab population who were not selected for positive family history or early age of onset and concurrently performed lpWGS and Whole-Exome Sequencing (WES) on each sample. We imputed germline variants using publicly available reference panels and assessed their accuracy using the paired WES samples. Using the imputed variants, we calculated a population-adjusted PRS and discovered various interactions between polygenic risk and other clinical features such as family history, pathogenic rare variant burden, and age of onset. Altogether, our investigation demonstrated an approach of using PRS beyond risk prediction to understand the polygenic risk landscape in an understudied population and highlighted the importance of studying diverse populations in expanding our knowledge about breast cancer.

## Methods

### Study Participants

Blood samples from 220 female breast cancer patients from the indigenous Arab population unselected for early age of onset or family history of cancer were collected from 2 participating institutions in Eastern Saudi Arabia: King Fahd Hospital - Alhafouf and King Fahd University Hospital - Dammam. This study was conducted under the following IRB protocol (IAU-IRB#2019-01-109) and conforms to the Declaration of Helsinki.

### Sequencing and Library Preparation

All samples prepared for lpWGS had sufficient starting material (100 ng of double-stranded gDNA). Normalized DNA was fragmented (Covaris sonication) to 350 bp and then ligated to specific adapters during automated library preparation (Roche/KAPA, Hyper KK8504) using the Beckman FXp liquid handling robot. Libraries were pooled in equal volume and sequenced on an Illumina nano flow cell to estimate each library’s concentration based on the number of index reads per sample. Library construction is considered successful if the yield is larger than or equal 250. All samples were successful. Libraries were normalized, pooled, and sequenced using Illumina platforms. Pooled samples were demultiplexed using the Picard tools version 1.130 [11].

For whole-exome sequencing (WES), a total amount of 1.0μg genomic DNA per sample was used as input material for the DNA sample preparation. Whole-exome capture libraries were generated using Agilent SureSelect Human All ExonV6 kit, and fragmentation was carried out by a hydrodynamic shearing system (Covaris, Massachusetts, USA) to generate 180-280bp fragments. Products were purified using the AMPure XP system (Beckman Coulter, Beverly, USA) and quantified using the Agilent high-sensitivity DNA assay on the Agilent Bioanalyzer 2100 system. The qualified libraries were fed into Illumina sequencers after pooling according to their effective concentration and expected data volume. All case samples had satisfactory effective read rates (> 97%) and error rates (< 0.03%) and are included in further analysis.

### Alignment

All raw sequencing data were uploaded to Terra (https://firecloud.terra.bio/), a collaborative cloud-computing platform utilized for genomic analyses, developed as part of the NCI Cloud Pilot program and supported by the Broad Institute [12]. Using Genome Analysis Toolkit (GATK) version 4.1.8.1 [13], all FASTQ files were first converted into unaligned Binary Alignment Map (uBAM) files, then aligned to the human reference genome b38 using BWA (version 0.7.15), as recommended by the GATK best practice workflows [14].

### Sequencing Coverage

The average sequencing coverage of all lpWGS and WES samples was calculated using the GATK’s (version 3.7) tool “DepthofCoverage”. A sample-wide mean coverage of 0.1X was considered the minimum acceptable coverage for lpWGS, and a 15X average coverage over exon intervals was considered the minimum acceptable coverage for WES.

### Whole-Exome Variant Calling

DeepVariant (version 1.0.0) [15], a deep learning-based variant calling method that has demonstrated to have superior performance at detecting pathogenic variants compared to the standard joint-genotyping approach [16,17], was used to call germline variants from WES data (docker image: gcr.io/deepvariant-docker/deepvariant:1.0.0). All variants annotated with “PASS” in the FILTER column of the VCF were deemed high-quality. Variants passing QC from all samples were then merged into one VCF file using GATK’s (version 3.7) tool “CombineVariants”. Subsequently, the ‘vt’ tool (version 3.13) was used on the cohort VCF file to normalize and decompose multiallelic variants.

### Functional and Clinical Annotation of Germline Variants

The cohort VCF File was annotated using Variant Effect Predictor (VEP, release 104.3) [18] with the publicly available GRCh38 cache file with a custom plug-in to include a recent “ClinVar” database release (accessed in June 2021). Using the tier criteria used by the Catalogue of Somatic Mutation in Cancer (COSMIC)[19], only variants in “germline tier 1” genes were considered. All detected variants were then classified into five categories: benign, likely benign, variants of unknown significance, likely pathogenic, and pathogenic, using the American College of Medical Genetics (ACMG) guidelines[20]. Variants classified as likely pathogenic or pathogenic are collectively referred to as pathogenic variants (PV).

### Low-pass Whole Genome Imputation

To obtain variant calls from lpWGS, GLIMPSE v1.1.1 [21] was used to perform genome-wide variant imputation. Following the recommended steps, the genome-wide genotype likelihood was first calculated on each sample using bcftools then separated into smaller genomic intervals before imputation. To maximize the number of variants imputed, we used Eagle v2.4.1 [22] to computationally phase the publicly available 1000 Genome [23] (1KG) WGS VCFs that was called using DeepVariant [24] (v1.0.0, GLnexus v1.2.7, GRCh38 reference), and used the output as the reference panels for imputation. After imputation was carried out on each genomic chunk, they were combined using the “GLIMPSE_ligate” command with default arguments, producing the final imputed VCF.

### Imputed Variant Quality Control

To assess variant imputation accuracy and to select a proper filtering threshold, the concordance of exonic variants was calculated based on the intersection of variants called both by DeepVariant using WES data and imputed by GLIMPSE using lpWGS. The “INFO” score outputted by GLIMPSE, which is a value that ranges from 0 to 1 where 1 indicates high confidence in the variant call, is referred to as the imputation quality score. The linear transformation of the posterior genotype probabilities generated by imputation, which is a number ranging from 0 to 2 where a number close to 1 indicates confidence in a single alternate allele at the location while a number close to 2 reflects confidence in having 2 alternate alleles at the location, is referred to as variant dosage. Variants were binned based on minor allele frequency, and the correlation between variant dosage and the number of alternate alleles (0, 1, or 2) called by DeepVariant, referred to as dosage correlation, was calculated within each bin for every sample. Allele frequencies were calculated based on allele counts in the cohort.

### Relatedness Inference

To control for confounding effects from related individuals, PLINK 1.9 [25] was first used to extract biallelic single nucleotide polymorphisms (SNPs) from the merged WES VCF file. Subsequently, LDAK 5.2 [26] was used to compute a kinship matrix assuming the LDAK-Thin heritability model with a correlation squared threshold of 0.98 and window size of 100 kilobases, as recommended (https://dougspeed.com/calculate-kinships/). Samples were then removed until no pairs have a kinship value greater than 0.125. Five samples were removed after this step.

### Polygenic Risk Score Calculation

To assess the clinical applicability of PRS, we adopted a similar PRS calculation methodology proposed by Hao L. et al [27] and curated the initial sets of PRS weights from “CancerPRSWeb” [28], a repository that contains PRS coefficients for major cancer traits derived from multiple large population databases such as the UK BioBank (UKB) [29], Michigan Genomics Initiative (MGI) [30], and GWAS Catalog [1]. To pick PRS sets most relevant to breast cancer, we selected “Breast Cancer [Female]” as the cancer trait and manually curated 20 sets of non-subtype specific weights which had validation performance in either MGI or UKB. The number of SNPs in the selected weights ranged from 79 to 1,120,410, and they were derived using various methods with different performances in UKB or MGI, as measured by area under the receiver-operator characteristic curve (AUC_population_). After downloading the associated weight file and metadata, SNPs with hg19 coordinates were lifted over to hg38 using the python liftover library [31] for downstream compatibility. For each set of PRS weights, we calculated the unadjusted raw PRS in PLINK 1.9[25] by using the “--score” command with the “score-no-mean-imputation” option enabled.

### PRS Population Stratification Adjustment

To obtain the genetic principal components (PCs) of every sample, we first merged the Arab breast cancer cohort WES VCF with the WES VCF from the 1000 Genomes Project [23]. The merged WES VCF was then loaded using Hail v0.2[32] and filtered for variants with allele frequency > 0.05 and p-value greater than 1e-6 from the Hardy-Weinberg Equilibrium test. LD-pruning was then performed on passing variants with greater than 0.1 correlation within a 1 million base pair window. The Hail function “hwe_normalized_pca” was then applied to the resulting set of common variants, and the top 10 PCs were kept for further analysis.

To create a population-adjusted PRS, an ordinary least square model was fitted using the top 10 PCs as features with the raw PRS as the output variable. The difference between the predicted PRS and raw PRS was then standardized, creating the population-adjusted, residualized PRS. This process was then repeated for every set of PRS weights, and the CancerPRSWeb ID of the PRS weights with the highest performance at detecting breast cancer in first-degree relatives (*AUC*_*family*_) as well as any degree relatives (AUC_family-any_) was “PRSWEB_PHECODE174.1_Onco-iCOGS-Overall-BRCA_LASSOSUM_MGI_20200608”.

### Statistical Analysis

Unless otherwise specified, all odds ratios, 95% confidence intervals, and p-values were computed based on the two-sided fisher’s exact test, as implemented in the exact2×2 R package [33] with the argument “minimum likelihood correction”. Confidence intervals of the area under the receiver-operator characteristic curves (AUC) were calculated based on the formulation by J. Hanley and B. McNeil [34]. Statistical diagrams were visualized using Seaborn v0.11.2 [35]. Statistical models were constructed using the python package “statsmodel” [36].

To evaluate the change in PRS AUC after removing pathogenic variant carriers from the cohort, the p-values were obtained by calculating the proportion of samples in which the AUC was lower after removing pathogenic variant carriers using 10,000 bootstrapped samples of the analysis cohort.

## Results

### Sample Characteristics

All samples met the minimum sequencing coverage cut-off, where the median genome-wide coverage for lpWGS was 1.3X (interquartile range [IQR] 1.25-1.36X, Figure 1a) and median exome-wide coverage for WES was 48.1X (IQR: 44.8 – 51.8X, Figure 1b). After removing related individuals, a total of 215 female breast cancer patients of Middle Eastern ancestry were included in the final analysis (Methods). The WES variant calls were merged with variant calls of the 1000 Genomes Project, and the first 10 genetic principal components were calculated. As expected, due to the lack of Middle Eastern ancestry representation from the 1000 Genomes Project the PCs of the cohort form a cluster distinct from the rest of the samples (Figure 1c, 1d). Among those whose clinical information was available (n=200), the mean age of presentation was 47.8 years (SD 10.1 years). The clinical characteristics of the breast cancer cases including subtype and staging stratified by family cancer history status can be found in Supplementary Table 1.

**Figure 1.**
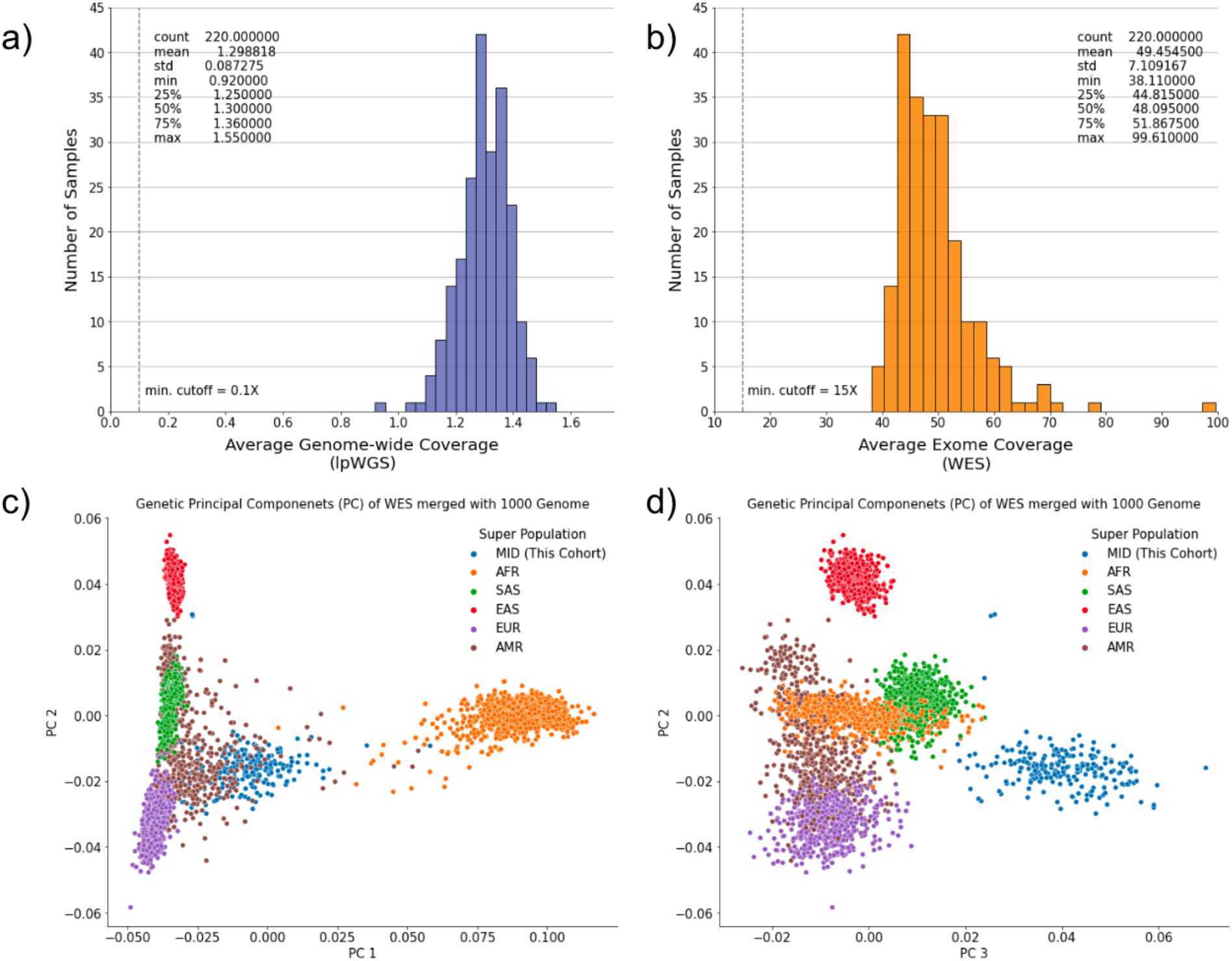
Sequencing metrics and sample characteristics of the cohort. a) The average genomewide coverage of lpWGS. b) The average exome coverage of WES. c & d) The first three genetic principal components based on the exomes merged with the 1000 Genomes data.

### lpWGS enables accurate imputation of low-frequency population-specific variants

To assess the quality of lpWGS-derived genotypes, we systematically analyzed the imputation performance of lpWGS in the exome-regions using high-coverage WES-derived high-quality variants as the ground truth. The median number of intersecting variants both called by WES and imputed by lpWGS per sample was 62,495 (IQR: 60,518-63,495), where at least 87.05% of the overlapping variants had imputation quality score greater than 0.8 across all samples (Figure 2a). To assess the reliability of the imputation quality score in reflecting the true posterior probability of the imputed variant having the specified variant dosage, we grouped imputed variants into bins by their imputation quality scores and calculated the dosages correlation for each sample within each bin (Methods). We observed good correspondence between imputation quality scores and genotype called from WES where variants in the 0.8-0.9 imputation quality score bin have median dosage correlation in a similar range (0.8441, IQR: 0.8355 - 0.8515) (Figure 2b, Supplementary Table S2). Collectively, the medians of dosage correlation per bin were highly positively correlated (Pearson correlation: 0.944, p < 0.001).

**Figure 2.**
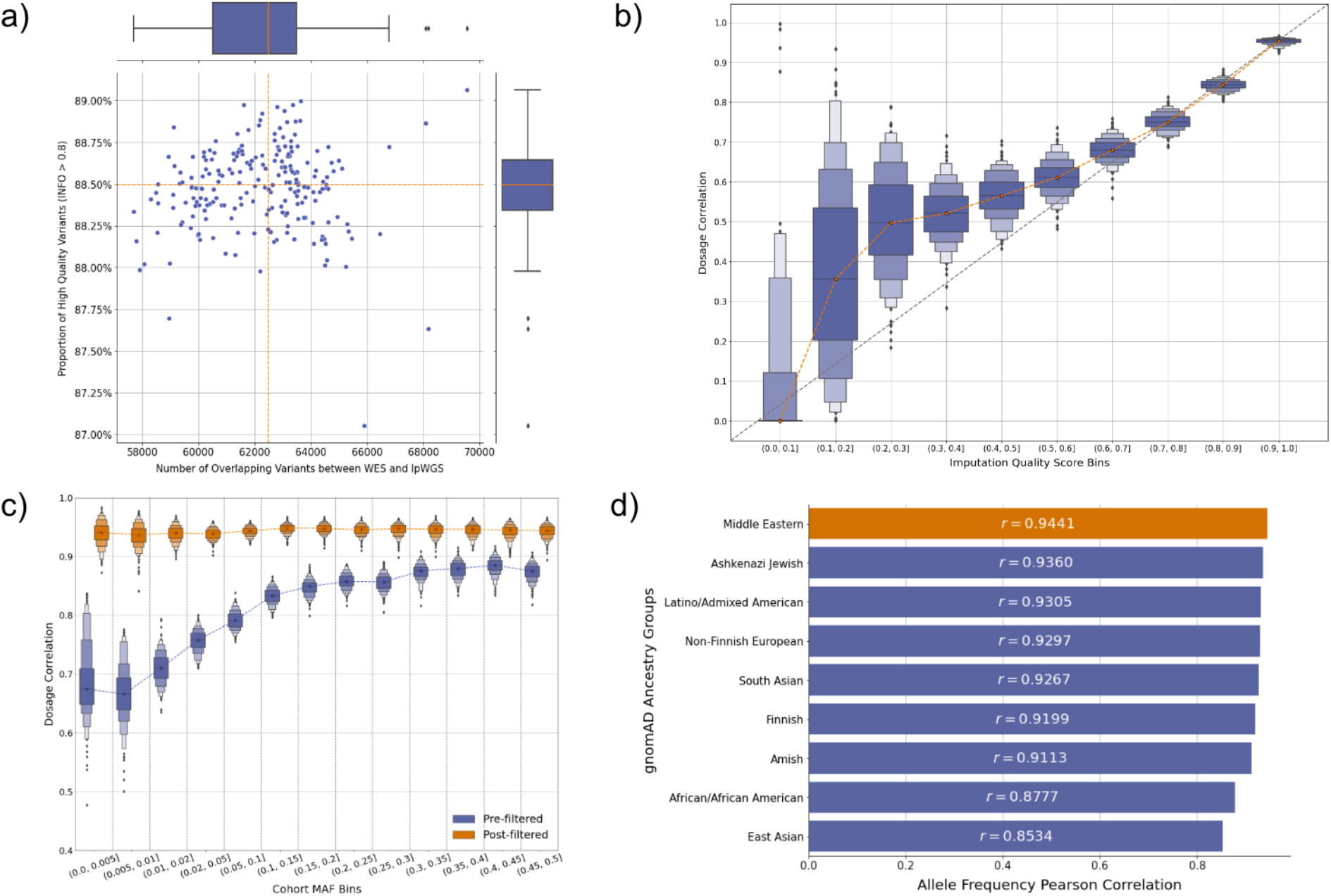
Imputation accuracy of lpWGS using high-coverage WES variants as ground truth. a) The number of variants both imputed by lpWGS and called by high-coverage WES per sample and the proportion of these variants with imputation quality score (INFO) at least 0.8. b) Boxen plot of the dosage correlation per sample grouped by imputation quality score intervals. Dosage correlation is defined as the correlation between the imputed variant dosage, which is a continuous value ranging from 0 to 2, and the number of alternate alleles called from WES c) The dosage correlation of lpWGS imputed variants grouped by cohort minor allele frequency before and after filtering out variants with imputation quality score below 0.8. We observe consistently strong performance after filtering regardless of MAF bins. d) The Pearson correlation between the allele frequency of the imputed variants in our cohort versus their allele frequencies in gnomAD ancestry groups.

Next, we evaluated the impact of minor allele frequency (MAF) on variant imputation quality by stratifying the variants into MAF bins and calculating the dosage correlation within each bin. We found after filtering out variants with imputation quality scores below 0.8, the dosage correlations are consistently strong regardless of MAF (Figure 2c, Supplementary Table S3), and collectively, the median dosage correlation per sample after filtering was 0.9459 (IQR: 0.9410-0.9490). As such, all variants with imputation quality score > 0.8 are included in downstream analysis without further filtering on MAF. To evaluate if the imputed variants are population-specific, we obtained the gnomAD [2] population allele frequencies for all imputed variants used for performance evaluation (n=284,601 variants) and calculated its Pearson correlation with the allele frequency in each of the gnomAD ancestry groups. As expected, we found our cohort’s variant minor allele frequencies to be the highest correlated with the Middle Eastern gnomAD ancestry group (Pearson correlation = 0.944, p < 0.001, Figure 2d).

### Imputed Variants Enables Calculation of Breast Cancer Burden Sensitive Polygenic Risk Score in the Arab population

Using the high-quality imputed variants of Arab breast cancer patients, we adopted a PRS calculation pipeline similar to the one proposed by Hao et al [27] and calculated 20 sets of breast cancer PRS for every sample using publicly available weights from “CancerPRSWeb”[28], a repository that contains PRS coefficients for major cancer traits derived from multiple large population databases. To account for population stratification, each PRS was residualized against the top 10 genetic principal components and then standardized for subsequent analysis (Methods, Supplementary Table S3). To evaluate each PRS’s ability to detect polygenic risk burden, we calculated the AUC of each PRS at the task of predicting patients with a self-reported family history of breast cancer at the first degree (AUC_family_). We found a positive correlation between the reported performance of the PRS at detecting breast cancer patients in larger, mostly European populations (AUC_population_) and AUC_family_ (spearman coefficient = 0.424, p-value = 0.0312) (Figure 3a), suggesting the calculated PRS was able to detect similar breast cancer burden from family cancer history as well as in general population.

**Figure 3.**
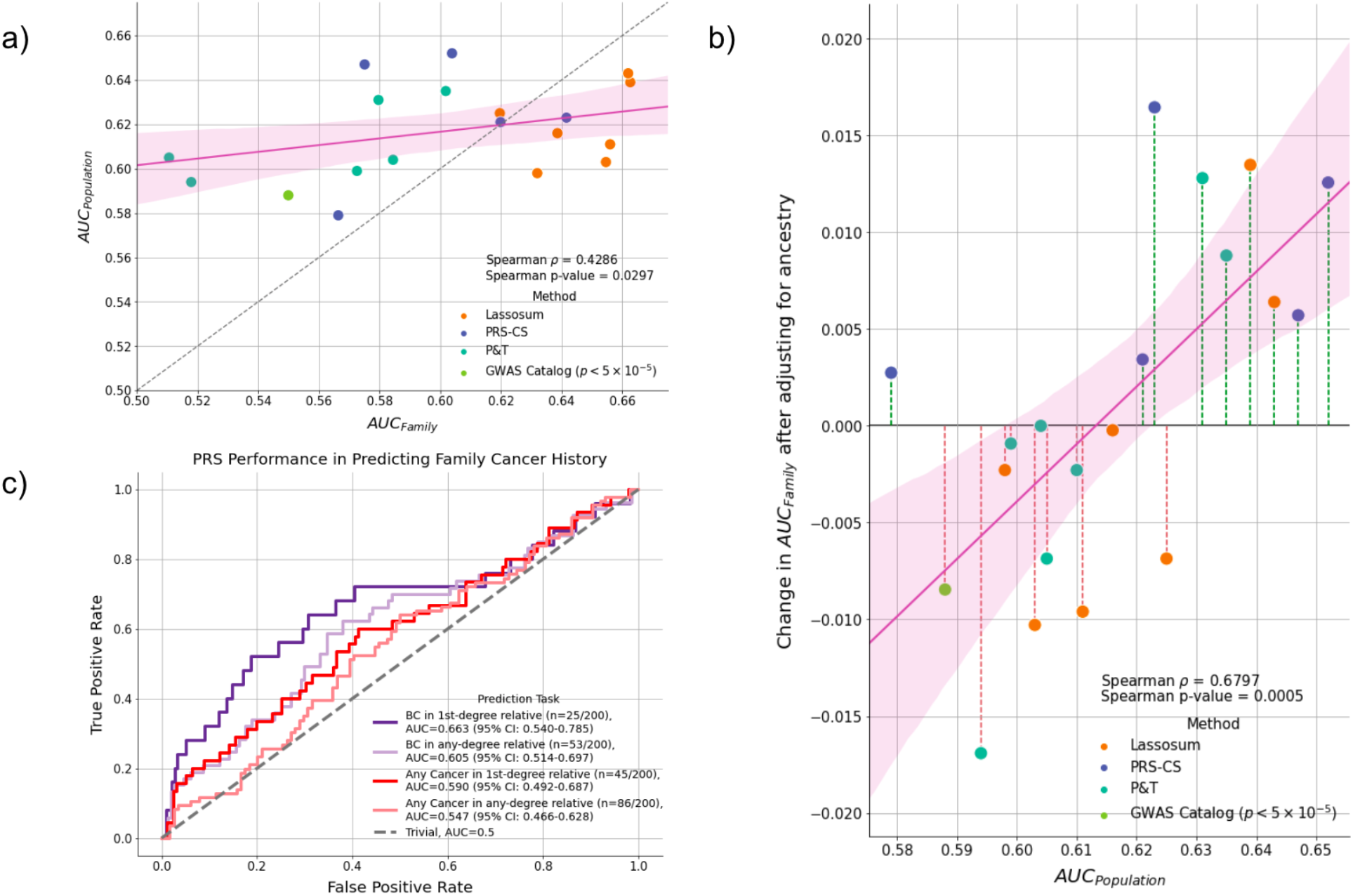
Evaluating the biological plausibility of the calculated PRS. a) The Spearman correlation between the performance of PRS at detecting breast cancer in first-degree relatives (AUC_family_) vs. the reported performance of the PRS at detecting breast cancer patients in larger European populations (AUC_population_). Method refers to the original method that was used to derive the weights for the PRS b) The original performance of the PRS (AUC_population_) plotted against the improvement in PRS AUC_family_ after the PRS is adjusted for population. The performance improvement is positively correlated with its original performance in the overall population suggesting the adjustment process was able to magnify burden effects while suppressing population stratification c) Evaluating the effectiveness of the best-performing PRS at predicting various cancer-related family histories. The PRS performs the best at predicting the presence of breast cancer in first-degree relatives and performance decreases as the relative degree increases and the cancer type become non-breast cancer-specific. BC: Breast Cancer.

To evaluate the effect of population adjustment on PRS, we compared the difference in AUC_family_ before and after applying residualization. We found the original population performance of the PRS to be positively correlated with the improvement in AUC_Family_ after adjusting for ancestry (Spearman correlation: 0.680, p-value: 0.0005) (Figure 3b). That is, for PRS with a lower AUC_poulation_, population adjustment resulted in lower performance in AUC_family_, while for PRS with a higher AUC_population_, population adjustment resulted in higher performance in AUC_family_. This suggests the population adjustment process was able to mask population-specific signals from PRS with lower AUC_population_ while amplifying causal signals from PRS with high AUC_population_.

Among the 20 sets of PRS for which the performance was evaluated (Supplementary Table S4), the PRS with the highest AUC_Family_ performance was chosen for downstream analysis (AUC_Family_: 0.663, AUC_Population_: 0.639, Number of SNPs: 118,388). To further validate the biological plausibility of the calculated PRS, we evaluated its performance at identifying patients with a family history of breast cancer or other cancers at varying degrees. We found the performance of the PRS to be the strongest at identifying patients with first-degree relatives with breast cancer (AUC_Family_: 0.663, 95%CI: 0.540-0.785), and the performance decreased when higher-degree relatives with breast cancer were included (AUC_Family-any_: 0.605, 95%CI: 0.514 - 0.697) or when non-breast cancer was included (AUC: 0.590, 95%CI: 0.492 - 0.687) (Figure 3c).

### Interaction of Mendelian and polygenic risk factors with family history in Arab breast cancer patients

Compared to patients with no first-degree relatives with breast cancer, Arab breast cancer patients who have first-degree relatives with breast cancer had higher PRS (p = 0.0086, two-sided Mann-Whitney U test (M.W.U.)) (Figure 4a). To understand if the burden of polygenic common variant risk is complementary to rare highly penetrant variant risk, we further stratified patients by rare germline pathogenic variant (PV) carrier status and performed association testing between each group. We found no significant difference in PRS distribution between PV carriers and non-carriers in patients with no first-degree relative with breast cancer (p = 0.946, M.W.U.), but among those who have a first-degree relative with breast cancer, non-PV carriers had significantly higher PRS than PV carriers (p = 0.0205, M.W.U.). In addition, among non-PV carrier patients, those with first-degree relatives with breast cancer have significantly higher PRS (p = 0.0002, M.W.U.) compared to those who do not (Figure 4a). In contrast, no difference in PRS distributions was found among PV carriers based on first-degree relative breast cancer status (p = 0.3142, M.W.U.).

**Figure 4.**
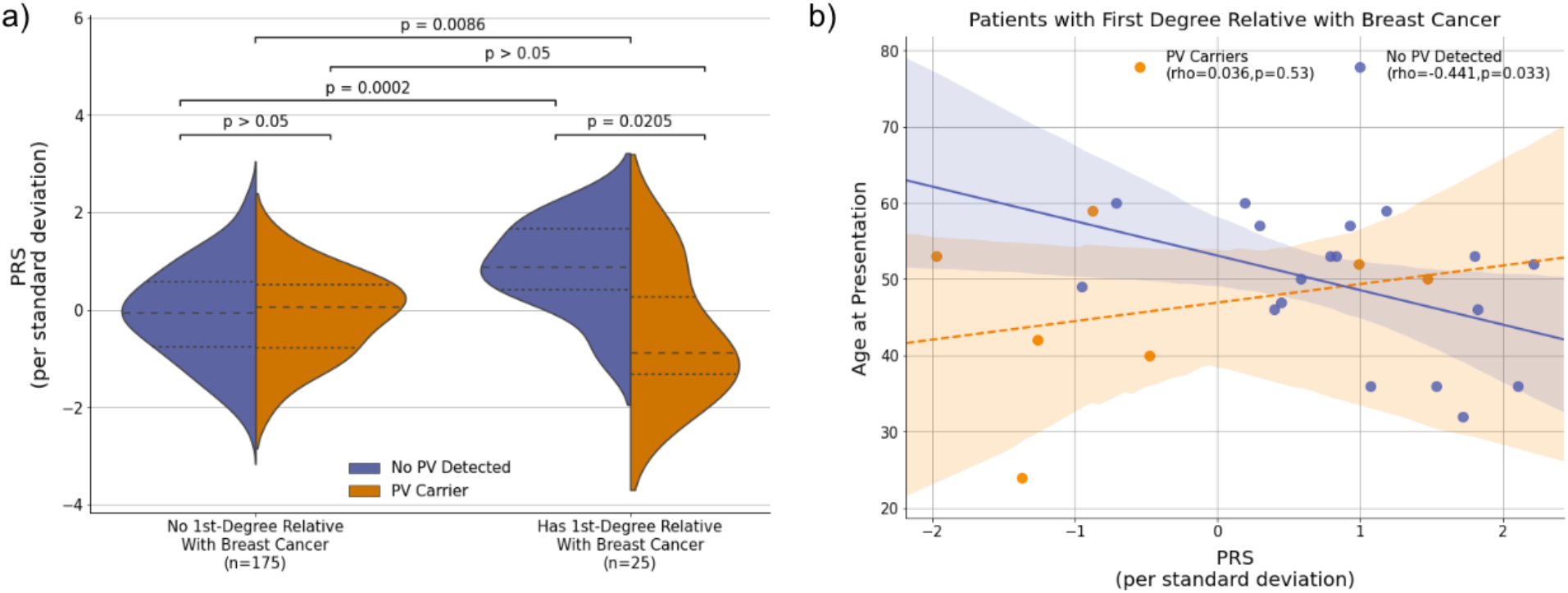
The interaction between rare pathogenic variant, PRS, and family breast cancer history in Arab patients with breast cancer. a) Violin plots of the distributions of PRS between patients with or without first-degree relatives diagnosed with breast cancer, stratified by rare pathogenic variant carrier status. The dotted line indicates the first and third quartile, the dashed line indicates the median b) Among patients with first-degree relatives diagnosed with breast cancer, age of onset is negatively correlated with PRS in patients with no detected pathogenic variants. PV: Pathogenic Variant

Given the performance of PRS at detecting familial breast cancer risk, we next assessed whether PRS could produce informative results for patients with a first-degree relative with breast cancer but are negative for rare germline pathogenic variants (n = 18). We found a statistically significant negative association between age of onset and PRS (Spearman rho: - 0.441, p = 0.033) (Figure 4b) among these patients, where each standard deviation increase in PRS corresponded to 4.52 (95% CI: 8.88-0.17, p = 0.042) years decrease in age of onset (intercept term: 53.09 years, 95%CI: 47.6-58.5, p < 0.001).

### PRS Performance is influenced by rare pathogenic variant carrier status

Given the detected interaction of PRS with rare pathogenic variants and age at diagnosis, we next investigated whether the performance of PRS is improved when PV carriers were removed from the cohort. We first stratified the cohort by PRS deciles and observed that compared to those in the middle deciles (Q2-Q9), patients in the top PRS decile are 5.53 (95% CI: 1.76-17.97, p = 0.003) times more likely to have a first-degree relative with breast cancer (Figure 5a). Upon removing PV carriers from the cohort, the bottom decile of PRS was depleted of any patients with first-degree relatives with breast cancer, and those in the top decile are now 7.34 (95% CI: 2.04-26.66, p = 0.002) times more likely to have a first-degree relative with breast cancer compared to those in the middle deciles (Figure 5b). A similar trend was seen for other groups, where the odds ratio of the top decile group having a family member with breast or other cancer compared to lower decile groups increased upon removing PV carriers. To systematically assess the impact of removing pathogenic variants from our cohort on the performance of PRS AUC, we reevaluated the performance of PRS at detecting relatives with breast cancer using 10,000 bootstrapped samples of the cohort. We calculated the p-value as the proportion of samples in which the AUC was lower after removing PV carriers and found that the removal of PV carriers leads to statistically significant (P < 0.05) increases in AUC performance across tasks (Figure 5c). In particular, the performance increased the most in detecting first-degree relatives with breast cancer where the difference in AUC was 0.11, or a 16.5% relative increase in AUC_Family_ after the removal of PV carriers. (Supplementary Table S5, Figure 5c).

**Figure 5.**
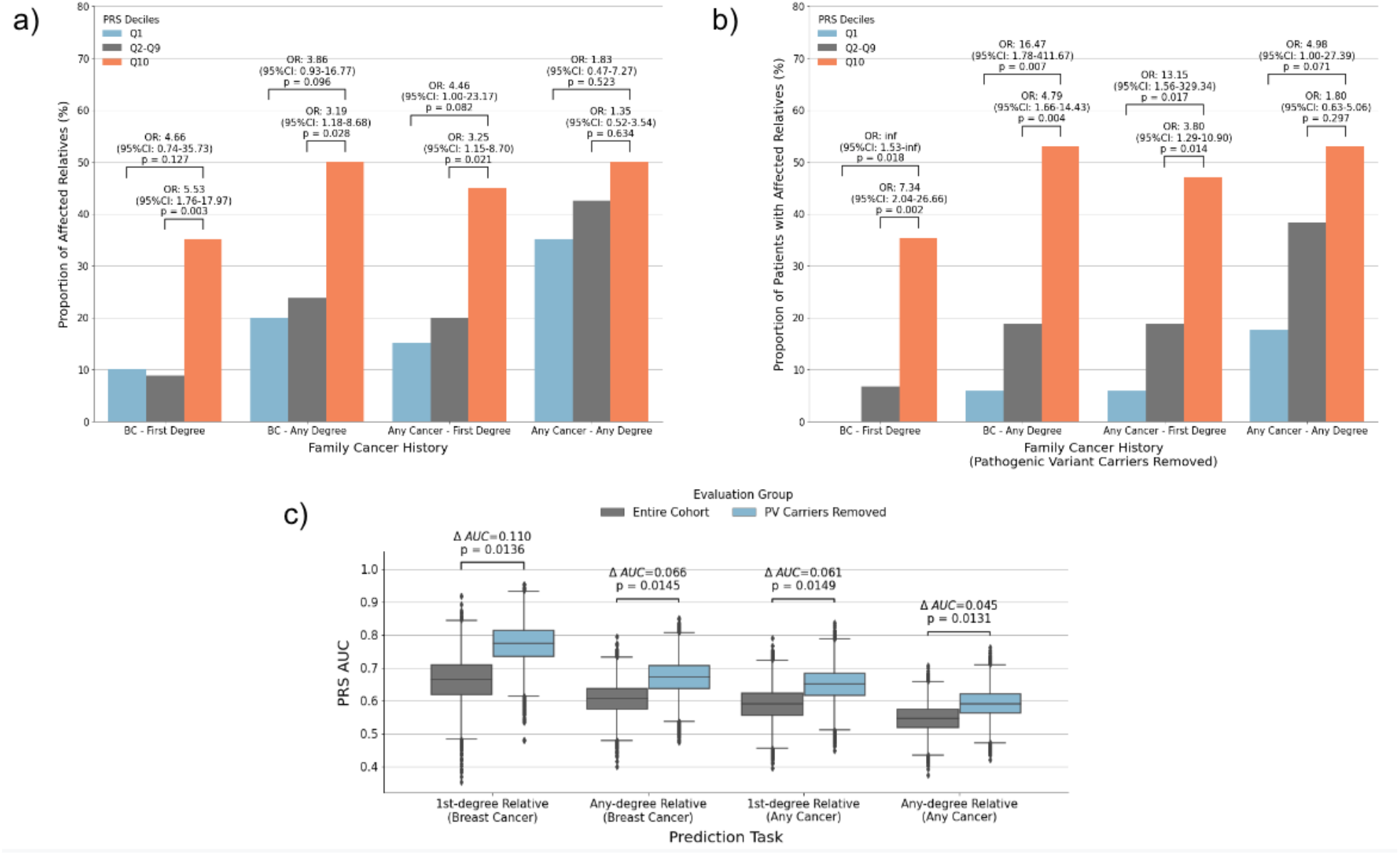
PRS Performance improves when pathogenic variants are accounted for. a) The proportion of patients in different PRS deciles having a family history of breast cancer or other cancers. b) The proportion of patients in different PRS deciles having a family history of breast cancer or other cancers after PV carriers are removed c) The performance of the PRS at detecting familial cancer risk before and after removing PV carriers from the cohort, as measured by AUC. P-value is based on the proportion of samples with lower AUC after removing PV carriers from 10,000 bootstrapped samples of the dataset. AUC delta refers to the difference in AUC after removing PV carriers in the original cohort.

## Discussion

Breast cancer is a global health burden, especially in understudied populations where the architecture of rare and common germline genetic determinants of the disease have been largely unexplored. In this multi-center study, we have shown that by utilizing lpWGS, we were able to impute high-quality population-specific variants as validated by WES. By adapting an existing PRS calculation pipeline proposed for clinical usage [27], we calculated a PRS that has the ability to detect breast cancer risk burden in the indigenous Arab populations and showed it to be negatively correlated with the age of presentation among patients with strong family history but have no detected rare pathogenic variant. This could have implications for current genetic screening guidelines, as individuals who qualify for genetic screening but have negative results from targeted gene panels may now have an additional way of assessing their genetic risks. In addition, we showed individuals with first-degree relatives with breast cancer have distinct PRS distributions based on PV carrier status, providing evidence for the genetic liability threshold model of breast cancer where the threshold for breast cancer may be achieved through a combination of rare or common variant risk [37,38]. Moreover, we showed the performance of PRS can be increased by accounting for rare pathogenic variant carrier status among patients. For future studies that may want to investigate the interactions between PRS and other clinical variables, this may be a useful strategy to employ to increase the power in detecting biological signals.

Overall, our study has also taken steps toward addressing disparities in genetics research in three ways. First, we have identified a set of PRS that performs well in detecting breast cancer burden among patients of Arab ancestry, who have traditionally been underrepresented in genomic studies. By showing the biological plausibility of the PRS, we made progress in creating a PRS whose clinical utility can be evaluated in this population. Second, we have demonstrated that lpWGS can be utilized to impute high-quality population-specific variants for an understudied population. Compared to high-coverage WGS, lpWGS is a more economical option both in terms of storage and computation. Because of this, lpWGS could be a viable strategy to rapidly increase data collection in understudied populations without compromising the types of analysis that can be performed. This is especially important in understudied populations where genotype data is scarce and budget may be a significant constraint.

Moreover, as methods improve, existing analysis pipelines on high-coverage WGS may eventually be applied to lpWGS data without substantial loss of power. Third, we demonstrated by using genetic principal components derived from diverse ancestries, the performance of PRS derived from pre-calculated weights can be further improved in understudied populations. This could increase the power of PRS association studies in understudied populations and serve as a quality-control step for investigating the transferability of existing PRS weights across ancestries.

As our understanding of how polygenic risk may affect breast cancer presentation expands, research focusing on incorporating such information in diverse populations is important. In this study, we have shown that a PRS that performs the best in detecting breast risk from the general population may not also be the best PRS at predicting familial breast cancer burden. In addition, some PRS have decreased performance when adjusted for the population while some improved. Understanding how to create PRS that is robust to population adjustment will be crucial to creating a PRS that is generally applicable to diverse populations. Moreover, while method developments are taking place in ensuring PRS has comparable risk prediction performance across ancestry [39], few have looked at the interactions between PRS with other clinical variables, especially in the context of understudied populations, which could be a missed opportunity to understand how ancestry-specific polygenic risk may affect disease presentation. Compared to cancer-predisposition variants that are under strong selection pressure, variants that do not affect fitness until certain phenotypes develop have less selection pressure and as a result may vary significantly across ancestries. By evaluating both the risk prediction capability of PRS and its ability to establish clinical correlations, we can reduce the likelihood of a “secondary disparity” scenario whereby even though a developed PRS is able to predict risk well across ancestries, it is unable to provide further clinical values such as predicting prognosis or responses to therapeutics in non-Europeans. Overall, understanding the potential utility of PRS in understudied populations is important to both addressing existing health inequalities and revealing novel biological insights.

## Limitations

This study has several limitations. First, due to the limited number of publicly available genomewide variant calls from individuals of Middle Eastern ancestry, the quality of the PRS is assessed indirectly using family cancer history instead of a case-control analysis. Second, while the imputation quality was satisfactory, a population-specific reference panel could further increase imputation quality. Third, the imputation performance was evaluated based on exome-only, and we assume the imputation performance would be similar in non-coding regions, which may not necessarily hold. Fourth, while this is one of the largest studies that investigated the polygenic risk of breast cancer in the GME region, the sample size is still relatively small compared to studies conducted on European populations, which may underpower our analysis. Finally, family histories of cancer are rarely fully reported, so some individuals with a negative family history may in fact have a family cancer history, further underpowering our analysis.

## Conclusion

Our multicenter observational analysis of 215 unrelated breast cancer patients identified a set of biologically plausible PRS capable of detecting breast cancer burden in the Arab population.This suggests lpWGS may be used as an effective tool complementary to high-coverage WES with both research and clinical values in understudied populations. We call for the expansion of genomic studies to be more diverse and for attention to PRS analysis in the context of the patients.

## Data Availability

All data produced in the present study are available upon reasonable request to the authors

## Data and Code Availability

All tools used in this study are publicly available. GLIMPSE is available at https://odelaneau.github.io/GLIMPSE/. PRS weights can be accessed on CancerPRSWeb [28]. The docker image containing DeepVariant is available at “gcr.io/deepvariant-docker/deepvariant:1.0.0” and the 1KG calls can be accessed through the google cloud bucket listed in the study: https://console.cloud.google.com/storage/browser/brain-genomics-public/research/cohort/1KGP/cohort_dv_glnexus_opt/v3. All data are available upon request.

## Acknowledgments

We thank all individuals who participated in this study. Dr. AlDubayan and Dr. Amein had full access to all the data in the study and took responsibility for the integrity of the data and the accuracy of the data analysis. This work was supported by King Abdulaziz City for Science and Technology, Riyadh, Saudi Arabia, grants #12-MED2226-46 and #11-MED2101-46, the Department of Defense Physician Research Award (W81XWH-21-1-0084, PC200150) (S.H.A), and the Department of Defense Idea Development Award - Early-Career Investigator (KC210042/W81XWH-22-1-0455) (S.H.A). The funding organizations were not responsible for the design and conduct of the study; collection, management, analysis, and interpretation of the data; preparation, review, or approval of the manuscript; and decision to submit the manuscript for publication.

## Authors’ contributions

[A. Al-Sulaiman, M. Al-Jumaan, Y AlMarzooq, F. Alhulhim, C. Vatte, A. Al Nemer, A. Almuhanna, A. Al-Ali] are responsible for the acquisition of clinical data and the enrollment of patients. [H. Chu, S. H. AlDubayan, S. Camp, S. Han] created the computational pipeline and processed the genetic data. [H. Chu, S. H. AlDubayan] performed analysis and interpretation of data. [H. Chu, S. H. AlDubayan] drafted the manuscript. [H. Chu] prepared the figures. [A. Al-Ali, S. H. AlDubayan, E. Van Allen, R. Gillani] performed critical revision of the manuscript for important intellectual content. All authors reviewed and edited the manuscript.

## Ethics Declaration

All individuals in this study consented to institutional review board-approved protocols that allowed for comprehensive genetic analysis of germline samples (Methods). This study conforms to the Declaration of Helsinki.

## Reference

1. Buniello A, MacArthur JAL, Cerezo M, Harris LW, Hayhurst J, Malangone C, et al. The NHGRI-EBI GWAS Catalog of published genome-wide association studies, targeted arrays and summary statistics 2019. Nucleic Acids Res. 2019;47:D1005–12.

2. Karczewski KJ, Francioli LC, Tiao G, Cummings BB, Alföldi J, Wang Q, et al. The mutational constraint spectrum quantified from variation in 141,456 humans. Nature. 2020;581:434–43.

3. Abou Tayoun AN, Rehm HL. Genetic variation in the Middle East-an opportunity to advance the human genetics field. Genome Med. 2020;12:116.

4. Scott EM, Halees A, Itan Y, Spencer EG, He Y, Azab MA, et al. Characterization of Greater Middle Eastern genetic variation for enhanced disease gene discovery. Nat Genet. 2016;48:1071–6.

5. Hashim MJ, Al-Shamsi FA, Al-Marzooqi NA, Al-Qasemi SS, Mokdad AH, Khan G. Burden of Breast Cancer in the Arab World: Findings from Global Burden of Disease, 2016. J Epidemiol Glob Health. 2018;8:54–8.

6. Saad M, Mokrab Y, Halabi N, Shan J, Razali R, Kunji K, et al. Genetic predisposition to cancer across people of different ancestries in Qatar: a population-based, cohort study. Lancet Oncol. 2022;23:341–52.

7. Martin AR, Atkinson EG, Chapman SB, Stevenson A, Stroud RE, Abebe T, et al. Low-coverage sequencing cost-effectively detects known and novel variation in underrepresented populations. Am J Hum Genet. 2021;108:656–68.

8. Li JH, Mazur CA, Berisa T, Pickrell JK. Low-pass sequencing increases the power of GWAS and decreases measurement error of polygenic risk scores compared to genotyping arrays. Genome Res. 2021;31:529–37.

9. Wasik K, Berisa T, Pickrell JK, Li JH, Fraser DJ, King K, et al. Comparing low-pass sequencing and genotyping for trait mapping in pharmacogenetics. BMC Genomics. 2021;22:197.

10. Homburger JR, Neben CL, Mishne G, Zhou AY, Kathiresan S, Khera AV. Low coverage whole genome sequencing enables accurate assessment of common variants and calculation of genome-wide polygenic scores. Genome Med. 2019;11:74.

11. Toolkit P. Picard toolkit. Broad Institute, Github Repository [Internet]. 2019; Available from: https://broadinstitute.github.io/picard/

12. Birger C, Hanna M, Salinas E, Neff J, Saksena G, Livitz D, et al. FireCloud, a scalable cloud-based platform for collaborative genome analysis: Strategies for reducing and controlling costs [Internet]. bioRxiv. 2017 [cited 2022 Feb 13]. p. 209494. Available from: https://www.biorxiv.org/content/10.1101/209494v1

13. Van der Auwera GA, O’Connor BD. Genomics in the Cloud: Using Docker, GATK, and WDL in Terra. “O’Reilly Media, Inc.”; 2020.

14. Data pre-processing for variant discovery [Internet]. GATK. [cited 2022 Oct 10]. Available from: https://gatk.broadinstitute.org/hc/en-us/articles/360035535912-Data-pre-processing-for-variant-discovery

15. Poplin R, Chang P-C, Alexander D, Schwartz S, Colthurst T, Ku A, et al. A universal SNP and small-indel variant caller using deep neural networks. Nat Biotechnol. 2018;36:983–7.

16. AlDubayan SH, Conway JR, Camp SY, Witkowski L, Kofman E, Reardon B, et al. Detection of Pathogenic Variants With Germline Genetic Testing Using Deep Learning vs Standard Methods in Patients With Prostate Cancer and Melanoma. JAMA. 2020;324:1957–69.

17. Camp SY, Kofman E, Reardon B, Moore ND, Al-Rubaish AM, Aljumaan M, et al. Evaluating the molecular diagnostic yield of joint genotyping-based approach for detecting rare germline pathogenic and putative loss-of-function variants. Genet Med. Elsevier BV; 2021;23:918–26.

18. McLaren W, Gil L, Hunt SE, Riat HS, Ritchie GRS, Thormann A, et al. The Ensembl Variant Effect Predictor. Genome Biol. 2016;17:122.

19. Tate JG, Bamford S, Jubb HC, Sondka Z, Beare DM, Bindal N, et al. COSMIC: the Catalogue Of Somatic Mutations In Cancer. Nucleic Acids Res. 2019;47:D941–7.

20. Richards S, Aziz N, Bale S, Bick D, Das S, Gastier-Foster J, et al. Standards and guidelines for the interpretation of sequence variants: a joint consensus recommendation of the American College of Medical Genetics and Genomics and the Association for Molecular Pathology. Genet Med. 2015;17:405–24.

21. Rubinacci S, Ribeiro DM, Hofmeister RJ, Delaneau O. Efficient phasing and imputation of low-coverage sequencing data using large reference panels. Nat Genet. 2021;53:120–6.

22. Loh P-R, Danecek P, Palamara PF, Fuchsberger C A Reshef Y K Finucane H, et al. Reference-based phasing using the Haplotype Reference Consortium panel. Nat Genet. 2016;48:1443–8.

23. 1000 Genomes Project Consortium, Auton A, Brooks LD, Durbin RM, Garrison EP, Kang HM, et al. A global reference for human genetic variation. Nature. 2015;526:68–74.

24. Yun T, Li H, Chang P-C, Lin MF, Carroll A, McLean CY. Accurate, scalable cohort variant calls using DeepVariant and GLnexus. Bioinformatics [Internet]. 2021; Available from: http://dx.doi.org/10.1093/bioinformatics/btaa1081

25. Purcell S, Neale B, Todd-Brown K, Thomas L, Ferreira MAR, Bender D, et al. PLINK: a tool set for whole-genome association and population-based linkage analyses. Am J Hum Genet. 2007;81:559–75.

26. Zhang Q, Privé F, Vilhjálmsson B, Speed D. Improved genetic prediction of complex traits from individual-level data or summary statistics. Nat Commun. 2021;12:4192.

27. Hao L, Kraft P, Berriz GF, Hynes ED, Koch C, Korategere V Kumar P, et al. Development of a clinical polygenic risk score assay and reporting workflow. Nat Med. 2022;28:1006–13.

28. Fritsche LG, Patil S, Beesley LJ, VandeHaar P, Salvatore M, Ma Y, et al. Cancer PRSweb: An Online Repository with Polygenic Risk Scores for Major Cancer Traits and Their Evaluation in Two Independent Biobanks. Am J Hum Genet. 2020;107:815–36.

29. Sudlow C, Gallacher J, Allen N, Beral V, Burton P, Danesh J, et al. UK biobank: an open access resource for identifying the causes of a wide range of complex diseases of middle and old age. PLoS Med. 2015;12:e1001779.

30. Zawistowski M, Fritsche LG, Pandit A, Vanderwerff B, Patil S, Schmidt EM, et al. The Michigan Genomics Initiative: a biobank linking genotypes and electronic clinical records in Michigan Medicine patients [Internet]. bioRxiv. 2021. Available from: http://medrxiv.org/lookup/doi/10.1101/2021.12.15.21267864

31. Liftover [Internet]. Available from: https://github.com/jeremymcrae/liftover

32. Hail 0.2.98 [Internet]. Available from: https://github.com/hail-is/hail/releases/tag/0.2.98

33. Fay MP. Confidence intervals that match Fisher’s exact or Blaker’s exact tests [Internet]. Biostatistics. 2010. p. 373–4. Available from: https://www.niaid.nih.gov/about/brb-staff-fay

34. Hanley JA, McNeil BJ. The meaning and use of the area under a receiver operating characteristic (ROC) curve. Radiology. 1982;143:29–36.

35. Waskom M. seaborn: statistical data visualization. J Open Source Softw. The Open Journal; 2021;6:3021.

36. Seabold S, Perktold J. Statsmodels: Econometric and statistical modeling with python. Proceedings of the 9th Python in Science Conference [Internet]. SciPy; 2010. Available from: https://conference.scipy.org/proceedings/scipy2010/seabold.html

37. Neale B. Liability threshold models [Internet]. Wiley StatsRef: Statistics Reference Online. Chichester, UK: John Wiley & Sons, Ltd; 2014. Available from: https://onlinelibrary.wiley.com/doi/10.1002/9781118445112.stat06439

38. Wray NR, Maier R. Genetic Basis of Complex Genetic Disease: The Contribution of Disease Heterogeneity to Missing Heritability. Current Epidemiology Reports. 2014;1:220–7.

39. Ruan Y, Lin Y-F, Feng Y-CA, Chen C-Y, Lam M, Guo Z, et al. Improving polygenic prediction in ancestrally diverse populations. Nat Genet. 2022;54:573–80.

